# Prospective Observational Study of Post Surgical Pain in Women undergoing Mastectomy Surgery: Post Surgical Pain After Mastectomy (PPAM) Study

**DOI:** 10.64898/2026.09.19.26363470

**Authors:** Uchenna O. Umeh, Hyung G. Park, Randy Cuevas, Hamleini Martinez, Raven Perez, Juan P. Cata, Jing Wang, Lisa V. Doan

**Affiliations:** Department of Anesthesiology, Critical Care and Pain Medicine, Hospital for Special Surgery, New York, NY, USA; Department of Anesthesiology, Weill Cornell Medicine, New York, NY, USA; Department of Population Health, NYU Grossman School of Medicine, New York, NY, USA; Department of Anesthesiology, Perioperative Care and Pain Medicine, NYU Grossman School of Medicine, New York, NY USA; Department of Anesthesiology and Perioperative Medicine, The University of Texas-MD Anderson Cancer Center, Houston, TX, USA; Department of Neuroscience and Physiology, NYU Grossman School of Medicine, New York, NY, USA

**Author notes:** **Address correspondence to:** Jing Wang MD PhD: Department of Anesthesiology, Perioperative Care and Pain Medicine, NYU Grossman School of Medicine, 550 1^st^ Ave, 5^th^ floor, New York, NY 10016. co-senior author (Jing Wang, Lisa V. Doan). **Data Availability Statement** Data are available from the corresponding author upon reasonable request.

**Keywords:** Chronic Postsurgical Pain, Post Mastectomy Pain Syndrome, Persistent Pain, Cancer Pain Breast Cancer

## Abstract

**Objectives:** Breast cancer survivors who have undergone mastectomy are at risk of developing Post Mastectomy Pain Syndrome (PMPS), and socio-demographic factors such as race, ethnicity, and neighborhood disadvantage have been posited as risk factors for the development of PMPS. This observational study assessed development of PMPS across different racial/ethnic and socio-demographic categories for women undergoing mastectomy.

**Methods:** This prospective, observational study included women between 18 and 85 years of age, undergoing mastectomy surgery. The primary outcomes were the Brief Pain Inventory (BPI) pain severity subscale and the presence of PMPS, defined as the binary indicator of BPI average item score > 0, at 3 months post-surgery. Outcomes were assessed across different racial/ethnic and socio-demographic categories. Associations were evaluated using covariate-adjusted robust and logistic regression.

**Results:** One hundred women were enrolled, and 89 completed the primary outcome assessment. Mean BPI pain severity at 3 months was higher in the non-White group (mean 0.99, SD 1.37) than in the White group (mean 0.60, SD 1.03; p=0.062). After multivariable adjustment, the difference was not statistically significant (regression coefficient for the White vs non-White group, -0.24; 95% CI, -0.78 to 0.19; p=0.263). PMPS was present in 51.5% of the non-White group and 35.1% of the White group. Adjusted odds of PMPS were lower in the White group but not statistically significant (OR 0.39, 95% CI 0.12 to1.17; p=0.099). BPI pain severity did not differ significantly by income, education, insurance type, or marital status.

**Conclusion:** This prospective observational study of pain after mastectomy surgery shows that there were no statistically significant adjusted differences in pain scores or the rate of development of PMPS at 3 months between White and non-White participants. The direction and uncertainty of the estimates support further investigation in larger, more diverse cohorts.

## 1. Introduction

Approximately 1 in 8 women (13%) will develop breast cancer over the course of their lifetime^1^. Breast cancer was the most common cancer in women in 157 out of 185 countries in 2022.^1^ Overall, Black women are more likely to die of breast cancer than women of any other racial or ethnic group^1^, with causes posited to include poor access to health care, low health literacy, and diagnosis at advanced stages of the disease.^2^

Advances in screening and improved treatment protocols have increased the number of patients who survive breast cancer and live disease-free or in remission for decades. However, cancer survivors who have undergone mastectomy with and without chemotherapy and radiation are at risk of developing chronic post-surgical pain (CPSP), which is defined by the International Association for the Study of Pain (IASP) as pain that develops after a surgical procedure and persists for more than 3 months.^3,4^ These persistent pain syndromes after breast surgery and other treatments, including radiation and chemotherapy, are termed Post Mastectomy Pain Syndrome (PMPS). The prevalence of PMPS has been estimated to be between 20-68%.^5,6,7^ Causes include nerve damage in the axilla and/or chest wall during the initial excision surgery and chemotherapy and/or radiation therapy.^8^ PMPS limits quality of life, impairs functional recovery, and contributes to chronic opioid use in post-mastectomy patients.

Social determinants of health such as socioeconomic status, neighborhood disadvantage, residential segregation, unemployment, racial discrimination, social support, and social network have been examined in relation to breast cancer incidence and play an important role in the stage at diagnosis and survival ^9^. Neighborhood concentrated disadvantage, a term that embodies clusters of poverty characteristics, is one of several factors that prominently contributes to racial breast cancer health disparities in American women.^10^ African American women develop more aggressive breast cancer features, such as triple-negative receptor status and more advanced histologic grade and tumor stage and suffer worse clinical outcomes than White women.^10^ Prior studies also suggest that race, ethnicity, and neighborhood disadvantage are risk factors for the development of PMPS, ^1,2^ compatible with the growing evidence on the association of racial and socio-demographic disparities with breast cancer treatment and survival.^11,12,13^ However, there remains only limited data on the role that ethnic/racial disparities and other social determinants of health play on the development PMPS, and prospective studies are particularly lacking.

The Post surgical Pain After Mastectomy (PPAM) study was an observational sub-study of a parent trial Ketamine Analgesia for Long-lasting Pain relief After Surgery (KALPAS), a multisite clinical trial which aimed to examine the effect of ketamine on the development of PMPS in 750 women undergoing breast cancer surgery.^14^ The PPAM study examined PMPS across different racial/ethnic and socio-demographic categories at three months for participants who declined or were ineligible to participate in KALPAS.

## 2. Methods

### Study Design/Setting

The PPAM observational prospective study was supported by the National Institutes of Health through the NIH HEAL initiative under the award number UH3CA261067. It was approved by the University of Utah Institutional Review Board (IRB# 00138959). The study was conducted at two enrolling sites: NYU Langone Tisch Kimmel and The University of Texas - MD Anderson Cancer Center. Enrollment began on April 20^th^, 2022, and data collection was completed on April 23, 2024. The study followed all applicable STROBE Guidelines. This study was not conducted with patient and public involvement. However, the study design was informed by existing research that included patient perspectives.

### Participants

#### Eligibility Criteria

Women aged 18–85 years who were eligible for, but did not consent to, the KALPAS study and were undergoing elective oncologic breast surgery, including unilateral or bilateral mastectomy, prophylactic mastectomy, with or without lymph node dissection or immediate/delayed reconstruction. Key exclusion criteria included non-English/Spanish speakers, participation in another interventional pain trial, distant metastatic disease, schizophrenia or psychosis history, ketamine allergy or sensitivity, pregnancy, planned bilateral or more extensive flap reconstruction, ASA physical status 4–6, and prior or ongoing hormone therapy for gender transition to male.

### Prescreening and Consenting Phase

Eligible patients were approached during presurgical visits or contacted remotely by phone or video. Interested patients were then contacted by the research team, and baseline surveys were completed after written informed consent was obtained.

### Study Assessments and Outcomes: Baseline and Three Months

The primary outcome, pain severity, was assessed using the Brief Pain Inventory (BPI, 0-10 scale) as the mean of 4 severity items: current, worst, least, and average pain over the past 24 hours. PMPS was defined as a BPI average score > 0 at 3 months.

Secondary outcomes included BPI average pain, worst pain, and pain interference; surgical-site pain measured by the Breast Cancer Pain Questionnaire; and anxiety and depressive symptoms measured by the PROMIS Anxiety and Depression Short Form 4a.

Additional exploratory outcomes included neuropathic symptoms (PROMIS Neuropathic scale), fatigue (PROMIS Fatigue), sleep quality and duration (PROMIS Sleep Disturbance), physical function (PROMIS physical function) and additional mood outcomes. Opioid use was collected from medical records and patient report. Participants were screened for substance abuse behaviors with the Tobacco, Alcohol, Prescription medication, and other Substances Tool (TAPS). Pain Catastrophizing Scale (PCS), Patient Health Questionnaire (PHQ), Generalized Anxiety Disorder (GAD) were used to measure additional anxiety and depression symptoms.

Surgical data and postoperative analgesic use were also collected. Data collection time points are summarized in Table 1.

**Table 1.** Study Objective, Demographic and Outcome Measures.

| **Procedures** | **Study Visit 1**<br>**Screening and Enrollment**<br>**(prior to surgery)** | **Study Visit 2**<br>**Day of Surgery**<br>**(POD 0)**<br>**(Perioperative)** | **Study Visit 3**<br>**3 months**<br>**(- 7 days / + 30 days)** |
| --- | --- | --- | --- |
| Informed consent | X |  |  |
| Review of inclusion/exclusion criteria | X |  |  |
| Demographics | X |  |  |
| Medical history and medication history | X |  |  |
| Brief Pain Inventory (BPI)<br>(assessing surgical site pain) | X |  | X |
| TAPS- Part 1 | X |  | X |
| PROMIS-neuropathic pain quality 5a |  |  | X |
| Breast Cancer Pain Questionnaire (BCPQ) |  |  | X |
| Patient Global Impression of Change (PGIC) |  |  | X |
| PROMIS Anxiety Short Form 4a | X |  | X |
| PROMIS Depression Short Form 4a | X |  | X |
| Pain Catastrophizing Scale (PCS) | X |  | X |
| Patient Health Questionnaire (PHQ)-2 | X |  | X |
| Generalized Anxiety Disorder (GAD)-2 | X |  | X |
| PROMIS sleep disturbance–short form 6a | X |  | X |
| Sleep duration | X |  | X |
| PROMIS fatigue–short form 7b daily | X |  | X |
| PROMIS-physical function–short form 6b | X |  | X |
| Patient reported analgesics |  |  | X |
| Surgical Data Form (from medical records) |  | X |  |

### Sample Size

For the power calculation for this prospective study, we assumed 33% non-White participants and 67% White participants and used a two-sample t-test allowing unequal variances to determine the minimum detectable effect size for differences between the two groups at 3 months, with 80% power and a two-sided significance level of 0.05. With the initially planned sample size of 425, the study was powered to detect an effect size of 0.35. With the revised sample size of N = 80 evaluable participants, the study was powered to detect a medium-to-large effect size of Cohen’s d = 0.68. To ensure 80 evaluable participants, we targeted enrollment of approximately 100 patients, accounting for an anticipated 80% retention rate at 3 months.

### Statistical Methods

We first assessed the normality of BPI pain severity scores at 3 months using the Shapiro–Wilk test. We used Wilcoxon rank-sum tests to evaluate unadjusted group differences in the primary outcome, BPI pain severity at 3 months, as well as in the change in BPI pain severity from baseline. Five main factors of interest were considered: (1) the White group (non-Hispanic White participants) vs the non-White group; (2) higher education (bachelor’s degree or higher) vs others; (3) employment (full- or part-time employment) vs others; (4) higher income (>$75,000) vs lower income; and (5) married vs others. For the unadjusted income comparison, the analysis was restricted to participants in the higher- and lower-income groups, excluding participants whose income was not reported. Participants whose income was not reported were retained as a separate category in the adjusted income models.

Multivariable regression models were used to evaluate the primary and secondary outcomes, adjusting for the baseline value of the corresponding outcome when available, patient age, BMI, enrollment site, breast cancer stage at baseline, and indicators of lymph node dissection and reconstruction for the current surgery.

Because of the presence of outliers and non-normality in most of the continuous outcome data, we used robust regression with Huber’s psi function.^16,17,18^ For statistical inference, we used a nonparametric bootstrap with 2,000 resamples, repeatedly refitting the robust regression model within each resample. The resulting bootstrap distributions of the regression coefficients were used to calculate percentile-based 95% confidence intervals and two-sided empirical p-values for group differences. For binary outcomes, logistic regression was performed, adjusting for the same set of covariates used for the continuous outcomes; adjusted odds ratios and corresponding 95% confidence intervals were reported. Each model was based on participants with available outcome and covariate data for that analysis. All analyses were conducted using R Statistical Software.^15^

## 3. Results

### 3.1. Descriptive statistics

100 women were enrolled into the study (Table 2), from April 20^th^, 2022 to April 23 2024. Patients scheduled for mastectomy surgery were assessed for eligibility (N= 204). Of them, 41 were excluded or not approached for consent. Of the patients who were approached for the study, 63 declined to participate. Of the 100 consented participant cohort, 89 participants completed the primary outcome assessment at 3 months, while 11 participants were lost to follow-up or withdrawn (Figure 1- FLOW Diagram).

**Figure 1:**
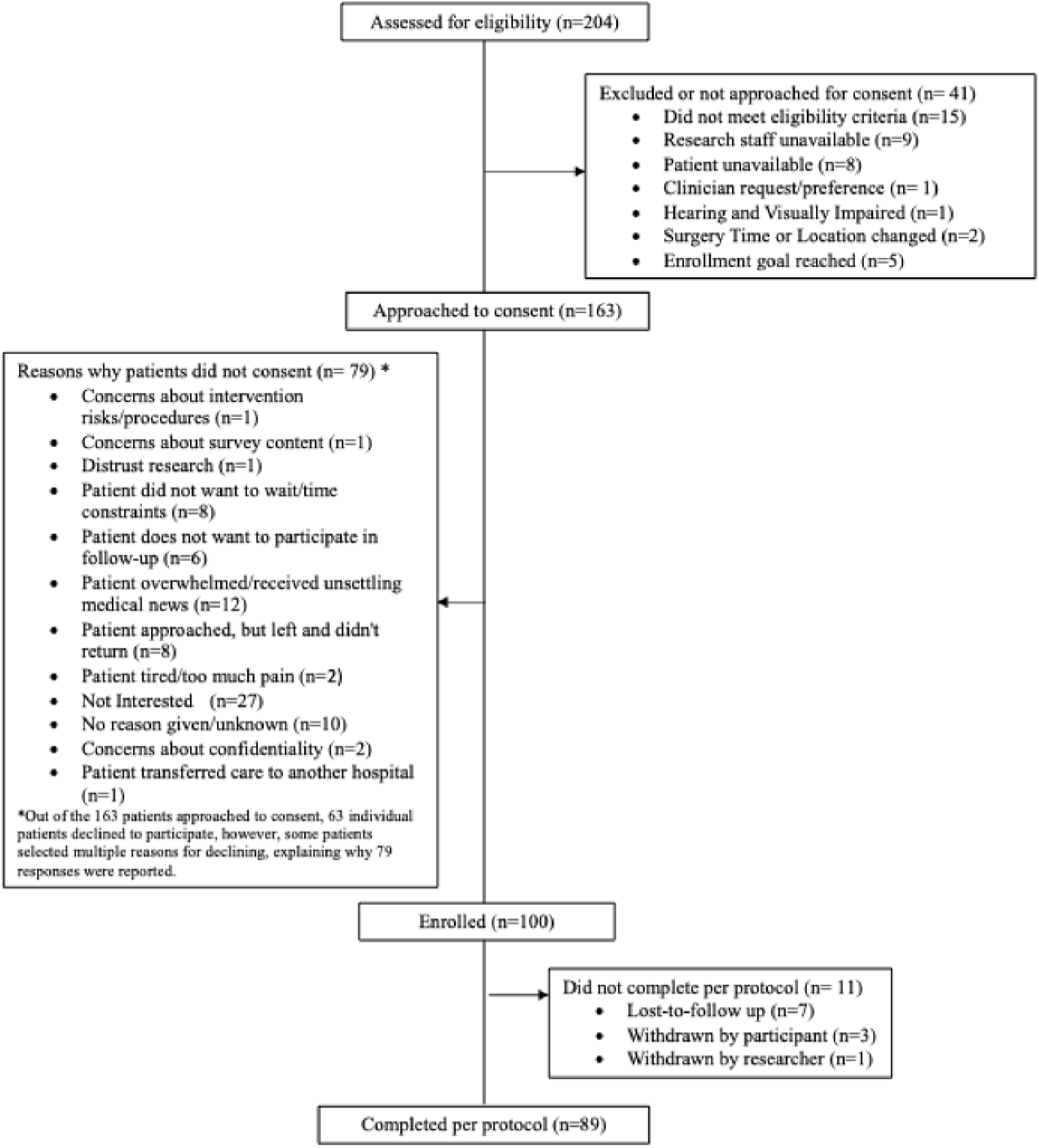
Flow Diagram.

**Table 2:**
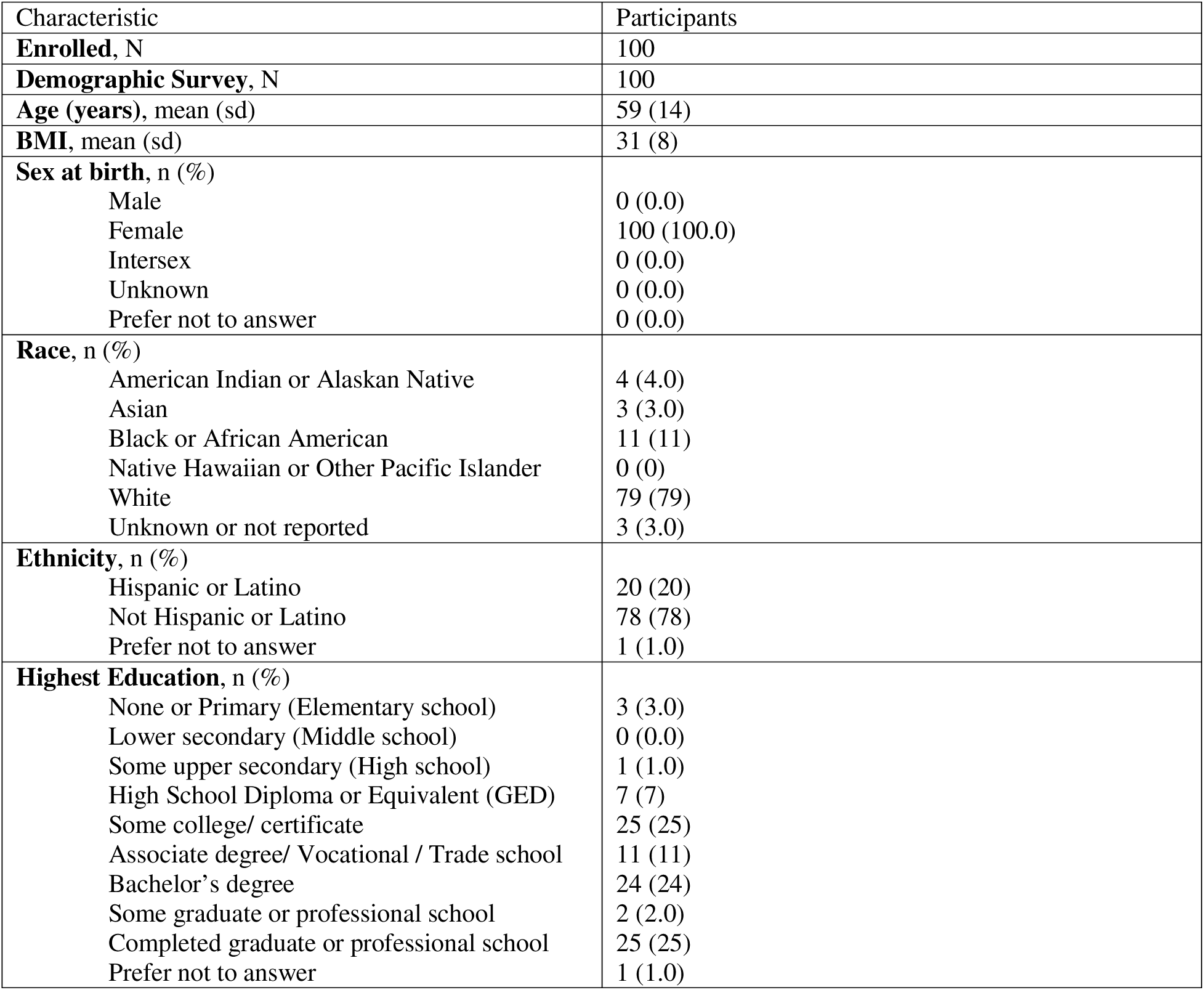
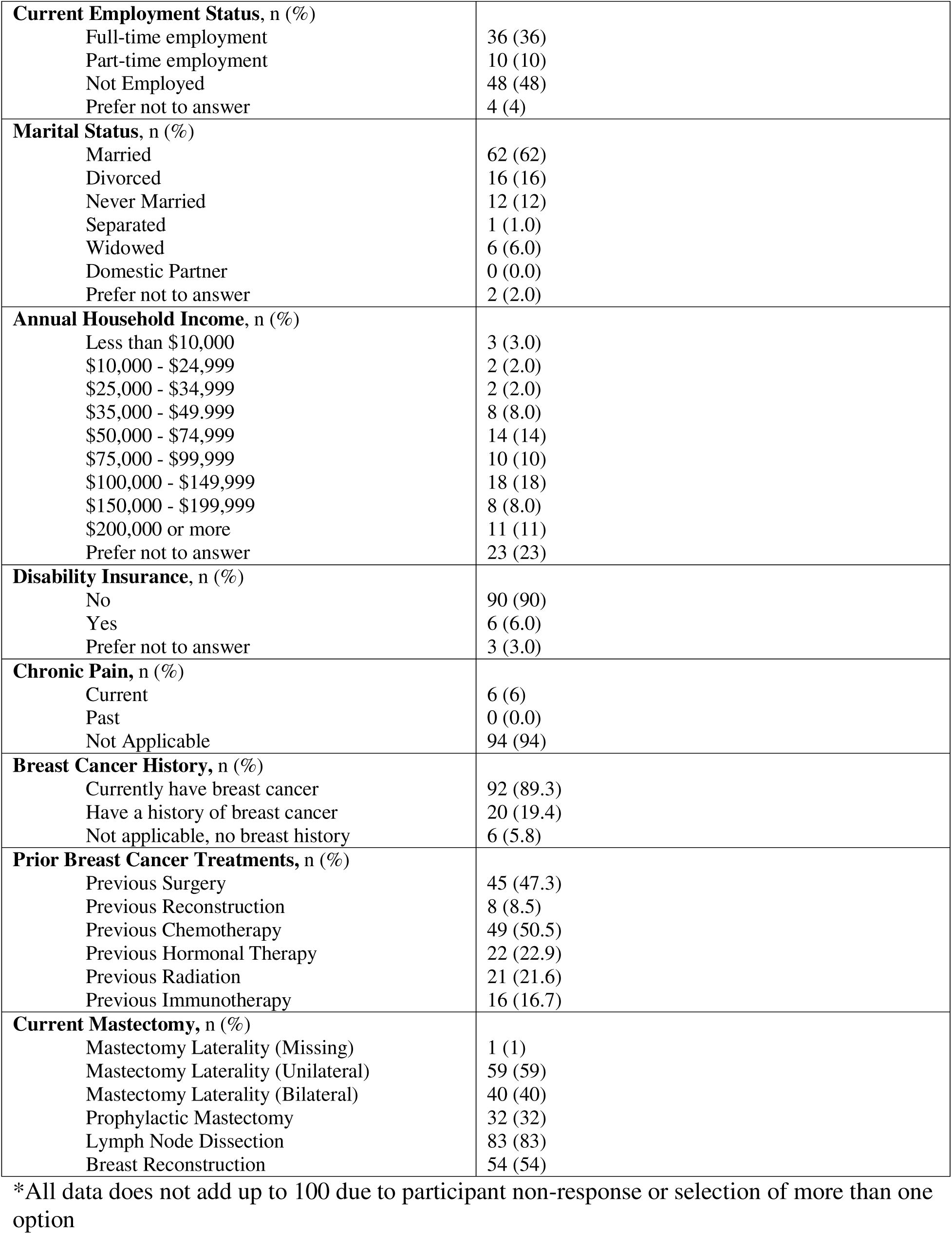
Demographics and Clinical Characteristics for PPAM Participants.

### 3.2. Association between PMPS and demographic factors

#### Race/ Ethnicity (Non-Hispanic White vs. Others)

The primary outcome – mean BPI pain severity at three months – did not differ statistically significantly between the White and non-White groups (mean: 0.99 (SD: 1.37) vs. 0.60 (SD: 1.03), P=0.063) (Figure 2). In multivariable regression adjusting for potential confounders, the estimated BPI pain severity score remained lower in the White group than in the non-White group, although the difference was not statistically significant (adjusted coefficient: −0.24; 95% CI: −0.78 to 0.19; P = 0.263).

**Figure 2:**
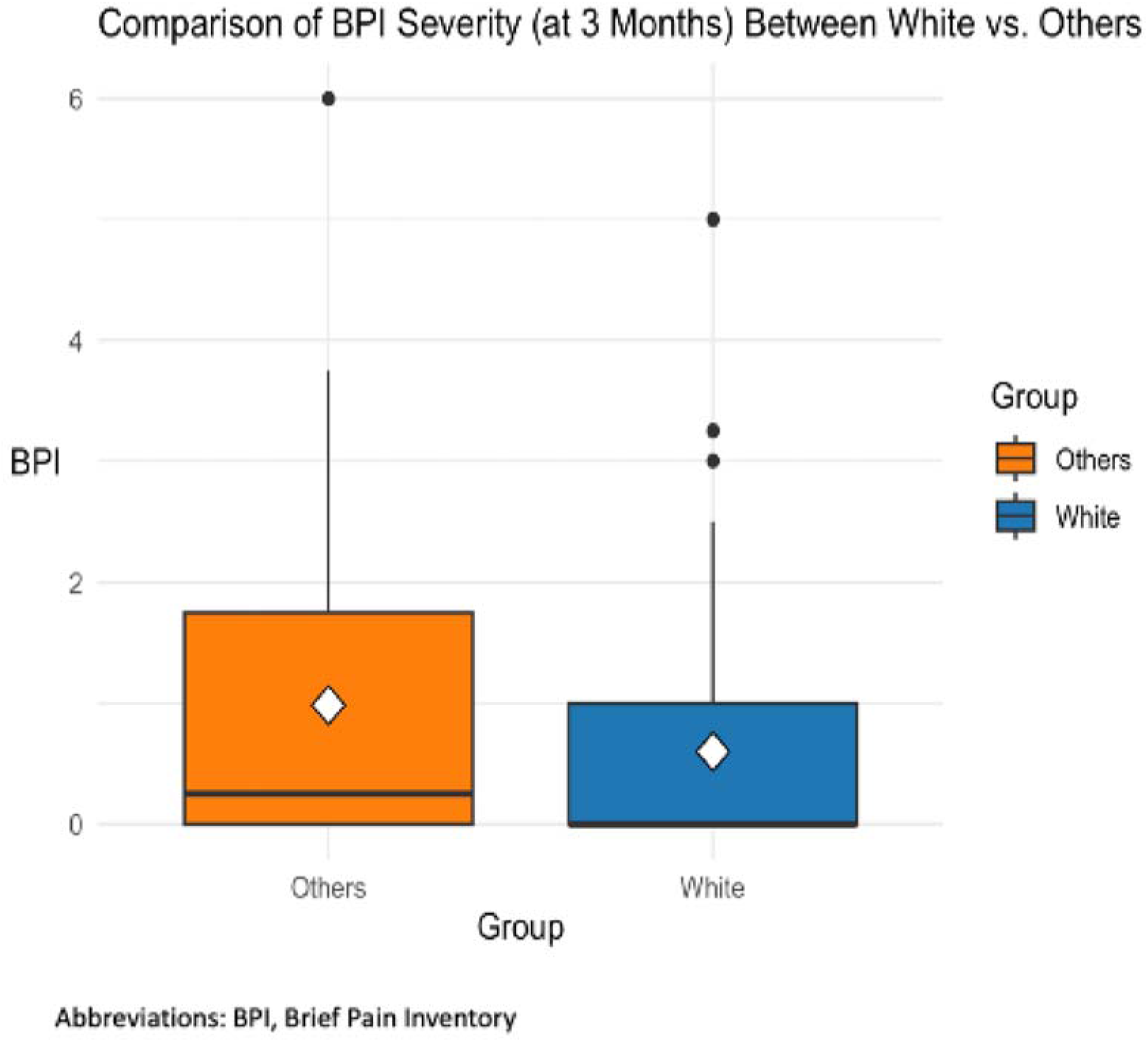
Boxplots of BPI severity at 3 months comparing White vs. All Others. The white diamond represents the mean, the horizontal black line indicates the median, boxes show the interquartile range (IQR), and whiskers extend up to 1.5×IQR.

We also examined the proportion of participants with PMPS, defined as a BPI average score >0, at 3 months. Among participants in the non-White group, 51.5% had PMPS compared with 35.1% of participants in the White group (Figure 3). In multivariable regression, the adjusted odds of having PMPS at 3 months were lower in the White group than in the non-White group (OR: 0.39; 95% CI: 0.12 to 1.17; P = 0.099), adjusting for baseline covariates and the baseline outcome. Similarly, the adjusted odds of having a BPI worst pain score >0 at 3 months were lower in the White group than in the non-White group (OR: 0.43; 95% CI: 0.14 to 1.28; P = 0.136). Although the confidence intervals included the null value, the direction of the estimates was consistent across these measures.

**Figure 3:**
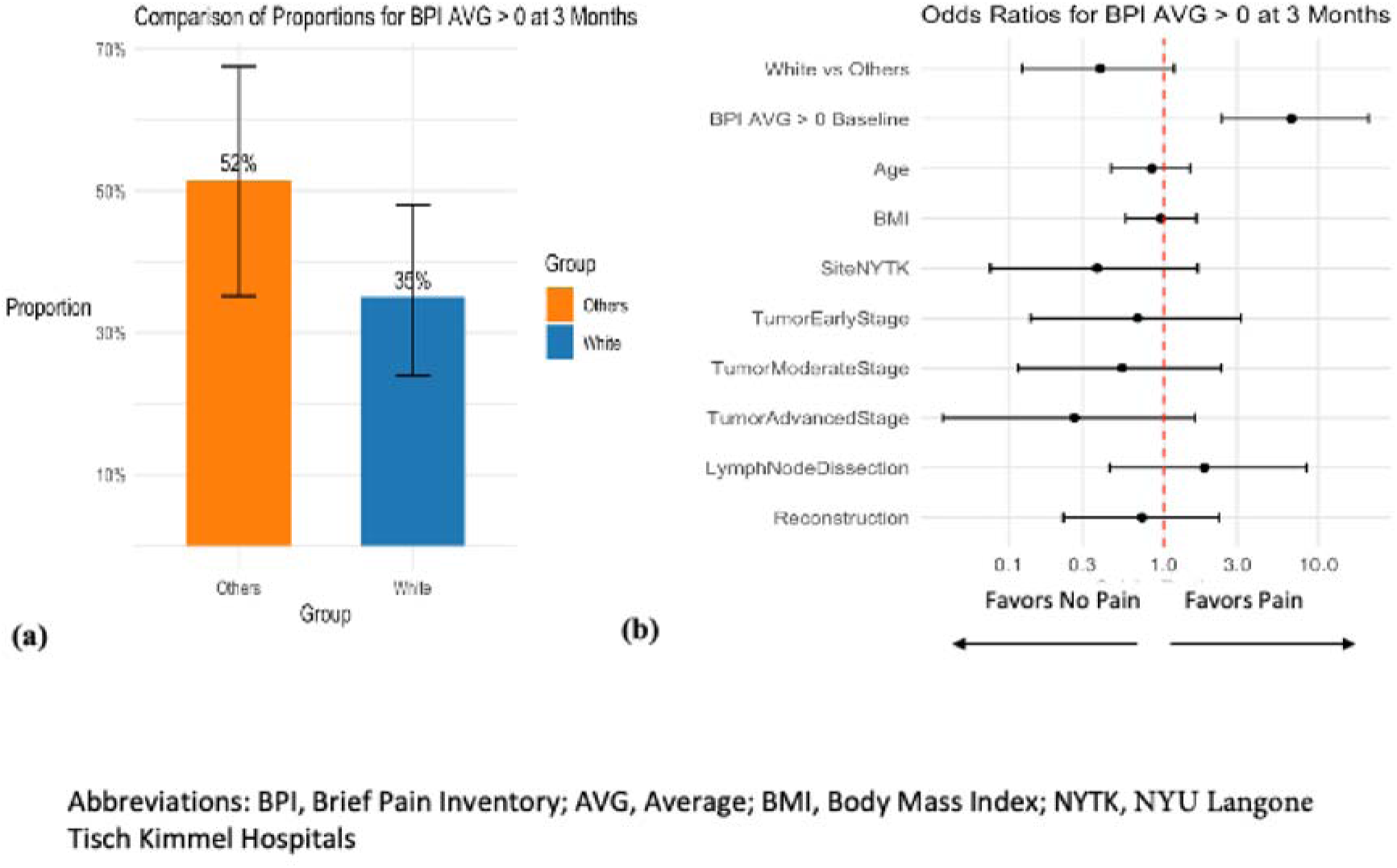
(a) Proportion of participants with BPI AVG > 0 at 3 months comparing White vs. All Others. Bars represent proportions with 95% confidence intervals. (b) Forest plot of odds ratios for BPI AVG > 0 at 3 months, adjusted for covariates. Points represent odds ratios with 95% confidence intervals. The dashed red line indicates OR = 1.

#### Education Level (Bachelor’s degree or Higher vs. Others)

There were no statistically significant differences in mean BPI pain severity at 3 months between participants with a bachelor’s degree or higher and other participants in either the unadjusted analysis (P = 0.534) or the multivariable analysis (adjusted coefficient: −0.11; 95% CI: −0.39 to 0.37; P = 0.767).

#### Employment Status (Full or Part Time Employment vs. Others)

There were no statistically significant differences in mean BPI pain severity at 3 months between participants with full- or part-time employment and other participants in either the unadjusted analysis (P = 0.341) or the multivariable analysis (adjusted coefficient: 0.26; 95% CI: −0.18 to 0.69; P = 0.255).

#### Income Level (Higher >$75,000 vs. Lower Income)

There were no statistically significant differences in mean BPI pain severity at 3 months between participants with annual income higher than $75,000 and those with lower income in either the unadjusted analysis (P = 0.790) or the multivariable analysis (adjusted coefficient: −0.08; 95% CI: −0.60 to 0.40; P = 0.684).

#### Marital Status (Married vs. Others)

There were no statistically significant differences in mean BPI pain severity at 3 months between married participants and other participants in either the unadjusted analysis (P = 0.835) or the multivariable analysis (adjusted coefficient: −0.26; 95% CI: −0.84 to 0.14; P = 0.199).

### 3.3. Association between secondary outcomes of pain and demographic factors

Multivariable analysis results, including robust linear regression coefficients with 95% confidence intervals for all available outcomes, along with reproducible code, are reported in Supplementary File, Section 5.

#### Other components of BPI scales

No statistically significant differences were observed across the demographic groups for the BPI average or BPI worst pain scores. Consistent with the primary outcome, adjusted scores were directionally lower in the White group than in the non-White group for both BPI average pain (adjusted coefficient: −0.42; 95% CI: −1.04 to 0.15; P = 0.146) and BPI worst pain (adjusted coefficient: −0.29; 95% CI: −1.01 to 0.39; P = 0.416), although the confidence intervals included zero. Race/ethnicity was also not statistically significantly associated with BPI interference (adjusted coefficient for the White group vs. the non-White group: −0.21; 95% CI: −0.96 to 0.32; P = 0.424).

#### Pain in Surgical Site

No statistically significant differences were observed between groups across all factors of interest as measured by the Breast Cancer Pain Questionnaire (BCPQ).

#### Anxiety

In multivariable analysis, higher income was associated with a higher PROMIS Anxiety Short Form 4a score compared with lower income (adjusted coefficient: 6.51; 95% CI: 2.05 to 11.19; P = 0.003). No other statistically significant differences in PROMIS anxiety scores were observed across the factors of interest.

#### Depression

No statistically significant differences were observed between groups across all factors of interest as measured by the PROMIS Depression Short Form 4a.

### 3.4. Association between exploratory outcomes of pain and demographic factors

#### Neuropathic symptoms, Fatigue, Sleep quality and duration, Physical function

No statistically significant differences were observed between groups across all factors of interest as measured by the PROMIS Neuropathic Quality 5a, PROMIS fatigue Short Form 7b, PROMIS sleep disturbance Short Form 6a, and PROMIS physical function Short Form 6b.

#### Additional Mood Outcomes

In multivariable analyses, higher income was associated with a lower Pain Catastrophizing Scale score compared with lower income (adjusted coefficient: −4.09; 95% CI: −7.04 to −0.83; P = 0.009). Higher education was also associated with a lower Patient Health Questionnaire-2 score compared with other education levels (adjusted coefficient: −0.40; 95% CI: −0.83 to −0.01; P = 0.040). No statistically significant differences were observed across the factors of interest for Generalized Anxiety Disorder-2 scores. These exploratory findings suggest that socioeconomic factors may relate differently to several aspects of postoperative psychosocial functioning and warrant confirmation in larger studies.

## 4. Discussion

Our study investigated the association between various socio-demographic factors and the development of persistent pain in breast cancer patients undergoing mastectomy surgery. The use of the BPI allowed us to quantify pain severity and pain interference and evaluate potential disparities across different socio-demographic characteristics.19 Although overall pain severity was low, approximately 41% of participants had a BPI average pain score >0 at 3 months, indicating that some degree of persistent pain was not uncommon. We did not identify statistically significant adjusted differences in pain outcomes between the White and non-White groups. Nevertheless, the non-White group had a descriptively higher mean BPI pain severity score than the White group (0.99 vs. 0.60) and a higher proportion of participants with PMPS (51.5% vs. 35.1%). The adjusted estimates were directionally consistent with these descriptive findings, although the confidence intervals were wide and included the null value. There were also no statistically significant differences in persistent pain across other characteristics, including educational level, employment status, income, or marital status.

Risk factors for development of persistent pain syndromes in breast cancer survivors can be divided into psychosocial, intraoperative and postoperative factors.^4^ Leyson et al.’s 2017 systematic review identified that BMI > 30, education < 12-13 years, lymphedema, not smoking, axillary lymph node dissection, chemotherapy, hormone therapy, and radiotherapy were significantly associated with higher odds for the development of chronic pain, with lymphedema being the biggest risk.^20^ In our study, lymph node dissection was not significantly associated with chronic pain development, although the odds ratio point estimate was greater than 1 and the confidence interval was wide (adjusted OR: 1.83; 95% CI: 0.45 to 8.39; P = 0.410) (Figure 3).

Other factors including smoking status were not examined. Among the breast cancer cohort, there is limited data on the role that race/ethnicity and other sociodemographic factors play on the development of persistent pain syndromes after mastectomy surgery. Previous studies have reported that non-White patients report more pain, decrease in physical functioning higher symptom intensity, and distress with breast cancer treatments compared to White women.^21,22,23^

Two prior studies showed notable differences from our results. Miaskowksi et al. examined risk factors for persistent pain after breast cancer surgery.^21^ Persistent pain was evaluated using the Breast Symptoms Questionnaire (BQS) prior to surgery and monthly for 6 months after surgery.^21^ The BQS Part 1 collected information on the occurrence of pain and other symptoms in the breast scar area. Those with pain the breast scar area completed Part 2 of the BSQ which rated the intensity of average and worst pain using the NRS scale.^21^ The authors showed that younger age, less education, being non-white, and lower total annual income are associated with reporting severe pain 6 months post-surgery.^21^ Another prospective study assessed socio-demographic, treatment-related, and health behavioral predictors of persistent pain 15 months and 7-9 years after breast cancer surgery.^24^ This study was conducted between 2001 and 2004, and assessed pain three months post-surgery in 1905 women.^24^ At 15-month post-surgery, 32.7% reported pain “almost every day” or more frequently and 20.4% at 7-9 years post-surgery.^25^ Socio-demographic factors including young age, lower education, lower income, lower occupational status, and health behavioral factors (smoking ≥ 10 cigarettes/day, obesity (BMI ≥ 30 and < 35), comorbidity, poor physical function) were significantly associated with pain at 15 months.^24^

Unlike the findings from both above studies, socioeconomic factors including employment status, marital status and annual income showed no significant differences in development of persistent pain at 3 months after mastectomy in our study. This could be attributed to two factors: first, our study followed patients for up to three months post-mastectomy, compared to 6 months in Miaskowski et al.’s study, and even longer--15 months and 7-9 years post-surgery--in the Johannsen et al. study. Second, overall symptom burden in our study appears to be low, as shown in the BPI scores. It should be noted that low symptom burden was also noted in a more recent trial on mastectomy.^25^ It is possible that contemporary surgical and anesthetic practices have decreased the overall PMPS severity. Finally, our study may have been underpowered due to a smaller sample size, particularly among patients from lower socioeconomic backgrounds, such as those who were unemployed, had education level less than a bachelor’s degree, or earned an annual income of less than $75,000.

Although socioeconomic characteristics were not significantly associated with BPI pain severity, the exploratory analyses identified associations with several psychosocial outcomes. Higher income was associated with higher PROMIS anxiety scores but lower pain catastrophizing scores, whereas higher education was associated with lower PHQ-2 scores. These findings suggest that socioeconomic characteristics may relate differently to specific dimensions of postoperative psychosocial functioning, even when differences in pain severity are not apparent. Given the exploratory nature of these analyses, these associations require confirmation in larger studies.

There are several limitations to this study. Study recruitment was limited to two urban academic medical centers, in New York City and Houston. Our study was likely underpowered. For instance, the observed effect size for the White vs. non-White group difference in mean BPI severity was quite small (Cohen’s d = 0.33) in this study. To achieve 80% power at alpha = 0.05, approximately 300 participants would have been needed. In addition, the non-White group combined participants from several racial and ethnic backgrounds, preventing evaluation of potentially important heterogeneity within this group. Future observational studies in this population can aim to recruit a larger sample size including more diverse and representative population and follow subjects beyond three months post mastectomy.

In conclusion, in this prospective observational study of persistent pain after mastectomy surgery, overall pain severity at 3 months was low, although a substantial proportion of participants reported some degree of persistent pain. Adjusted differences in pain severity and the occurrence of PMPS between the White and non-White groups were not statistically significant. However, pain severity and PMPS prevalence were consistently higher in the non-White group, and the confidence intervals remained compatible with potentially meaningful group differences. Other socio-demographic characteristics, including income below $75,000 per year, less than a college education, and unemployment, were not significantly associated with persistent pain. The observed estimates and exploratory psychosocial findings provide useful prospective data to inform larger and longer-term studies of disparities in pain and recovery after mastectomy.

## Data Availability

Data are available from the corresponding author upon reasonable request.

## CREDIT AUTHOR STATEMENT

**Uchenna O. Umeh**: Conceptualization, Methodology, Writing- Original Draft, Writing- Review and Editing. **Hyung G. Park**: Conceptualization, Methodology, Formal Analysis, Writing- Original Draft, Writing- Review and Editing. **Randy Cuevas**: Investigation, Project Administration. **Hamleini Martinez**: Investigation, Project Administration. **Raven Perez**: Investigation, Project Administration. **Juan P. Cata**: Methodology, Investigation, Writing- Review and Editing. **Jing Wang**: Conceptualization, Methodology, Supervision. **Lisa V. Doan:** Conceptualization, Methodology, Writing- Original Draft, Writing- Review and Editing, Supervision.

## PREVIOUS PRESENTATION

This paper was presented at the American Society of Regional Anesthesiology and Pain Medicine 2025 Spring Meeting and accepted as an abstract to the American Society of Anesthesiologists’ 2025 Annual Meeting.

